# Evaluation of DNA-poli: study protocol of a randomised controlled trial to assess a digital platform for family cascade genetic testing and predictive genetic counselling

**DOI:** 10.1101/2024.11.04.24316732

**Authors:** Marlies N. van Lingen, Janine V. Meulenkamp, Marten A. Siemelink, Tessa C. Beinema, Randy Klaassen, Dirk K.J. Heylen, J. Peter van Tintelen, Lieke M. van den Heuvel

## Abstract

**Introduction:** The present uptake of predictive genetic counselling among at-risk relatives (ARRs) for cardiogenetic diseases is suboptimal with 40-50% of ARRs being tested after one to three years post-disclosure. Digital technologies are increasingly proposed to improve accessibility, efficiency, and uptake of predictive genetic counselling and, if desired, predictive genetic testing. Therefore, DNA- poli was developed: a digital platform providing family communication support and pre- and post-test genetic counselling for ARRs. The online DNA-poli aims to decrease the threshold for ARRs to seek genetic counselling without compromising the quality of care while increasing the efficiency of genetic care. Here, we describe the study protocol for a randomised controlled trial evaluating DNA- poli in clinical practice.

**Methods and analysis:** A non-inferiority multicentre randomised controlled trial with parallel-group design will be conducted. The intervention group using the DNA-poli platform will be compared to a control group receiving regular counselling. Probands with hypertrophic or dilated cardiomyopathy in whom a (likely) pathogenic variant in specific genes with definitive gene-disease validity is identified, will be included like their ARRs and physicians. The primary outcome is the uptake of cardiogenetic counselling six months post-disclosure with an extended follow-up of one year and stakeholders’ experiences. Secondary outcomes are informed decision-making in ARRs, empowerment, and the satisfaction of all stakeholder groups. In addition, the efficiency of consultations and the genetic care process will be analysed. Descriptive and inferential statistics will be performed to analyse data.

**Ethics and dissemination:** This study protocol was exempted from approval by the Medical Ethical Committee NedMec because the Act of Medical Research Involving Human Subject (WMO) was not applicable (no. 23-066/C). Study findings will be shared with stakeholders, published in journals, and will be presented at both international and national conferences.

Registration details NCT06431425 ClinicalTrials.gov

## INTRODUCTION

Pathogenic and likely pathogenic gene variants causing inherited cardiac conditions (ICCs) can have major consequences if they remain unidentified [1]. These variants are characterised by incomplete penetrance and variable expression leading to a diverse phenotype that manifests in a wide age range [2 ,3]. ICC-associated (likely) pathogenic variants are generally inherited in an autosomal dominant fashion, indicating a 50% chance of inheritance for first-degree relatives [1]. Effective clinical and lifestyle interventions are available to reduce the morbidity and mortality of ICCs [4].

Therefore, if a (likely) pathogenic variant is known to underlie the phenotype in the proband, predictive genetic testing is offered to first-degree relatives [1]. It is performed stepwise, also known as cascade genetic testing [5 ,6]. Predictive testing of at-risk relatives (ARR) enables targeted clinical evaluation and subsequent risk stratification [4].

In current practice, a family-mediated approach is used to perform cascade genetic testing. Probands are asked to inform ARRs, generally supported by a family letter from a genetic counsellor [7 ,8]. Despite efforts to support probands in informing ARRs, the present uptake of genetic counselling for predictive DNA testing in ICC is relatively low, i.e., well below 50% one year after identifying the causative variant in the proband [8–10]. A retrospective cohort study in hypertrophic cardiomyopathy (HCM) performed by van den Heuvel et al. reports an uptake of genetic counselling of 60% after 16 years [9]. Furthermore, there is an increasing need for genetic counselling services due to the growing number of patients [11]. At the same time, limited numbers of genetic counsellors and resources result in long waiting lists and increased pressure on genetic care services [11].

Digital tools have therefore been suggested as a promising solution for improving the uptake, efficiency, and accessibility of genetic counselling [12–15]. Adam et al. showed that an interactive online decision-support tool can be as effective in increasing knowledge as current clinical practice [16]. This tool was well received as an addition, or even a replacement by the majority of the participants emphasising the feasibility of integration of such technologies into genetic care[16]. Torr et al. also designed a digital pathway for genetic counselling of patients with inherited breast cancer, resulting in increased uptake and high participant satisfaction[17]. In addition, a systematic literature review by Lee et al. found that digital tools generally improve knowledge and contribute to the decision-making process of counselees on genetic testing[14].

However, previously tested tools only supported pre-test counselling for probands [15] or were mainly focused on oncology [14]. Therefore, we propose “DNA-poli” (“poli” is Dutch for outpatient clinic), a digital informative platform providing pre- and post-test genetic counselling for ARRs in ICC. DNA-poli aims to decrease the threshold for ARRs to seek genetic counselling and increase the uptake while retaining counselling quality. Here, we describe the study protocol for a multicentre non-inferiority randomised controlled trial (RCT), in which DNA-poli is evaluated in HCM and dilated cardiomyopathy (DCM) probands, family members and genetic healthcare professionals (HCP) in a parallel group design.

### Setting and intervention

After a patient is diagnosed with HCM or DCM in clinical care, genetic testing is performed. If a likely pathogenic or pathogenic variant is identified, cascade genetic testing and pre-test genetic counselling are recommended for first-degree ARRs, i.e., biological parents, children, and siblings [4]. If a first-degree relative is deceased, second-degree ARRs should also be considered for pre-test counselling and cascade testing [4]. In current clinical care, a family letter is used to support probands with informing their relatives, accompanied by a general practitioner (GP) referral form for clinical and genetic evaluation and follow-up.

In DNA-poli, an online platform for family communication and pre-and post-test genetic counselling, most aspects of the cascade testing journey are digitalised. Functionalities and development of DNA-poli will be published elsewhere. After a secure login, probands can invite their ARRs with their permission to the online platform. Here, a concise and digital version of the family letter is available which is supported with illustrations. Referral by a GP is not necessary as DNA-poli involves research consultations. Next, after consenting to the DNA-poli pathway, ARRs can follow the pre-test journey in their own time and pace from wherever they like (Figure 1a). They can pause and leave at any time and return to continue where they left off. The pre-test counselling consists of four stages, (1) information provision on HCM or DCM, (2) heritability and genetic diseases, (3) genetic testing, and (4) decision support. In each stage, a virtual assistant with a static female embodiment called “Anna” is available to answer remaining questions and collect medical history via a pre-scripted chat conversation (Figure 1b).

**Figure 1.**
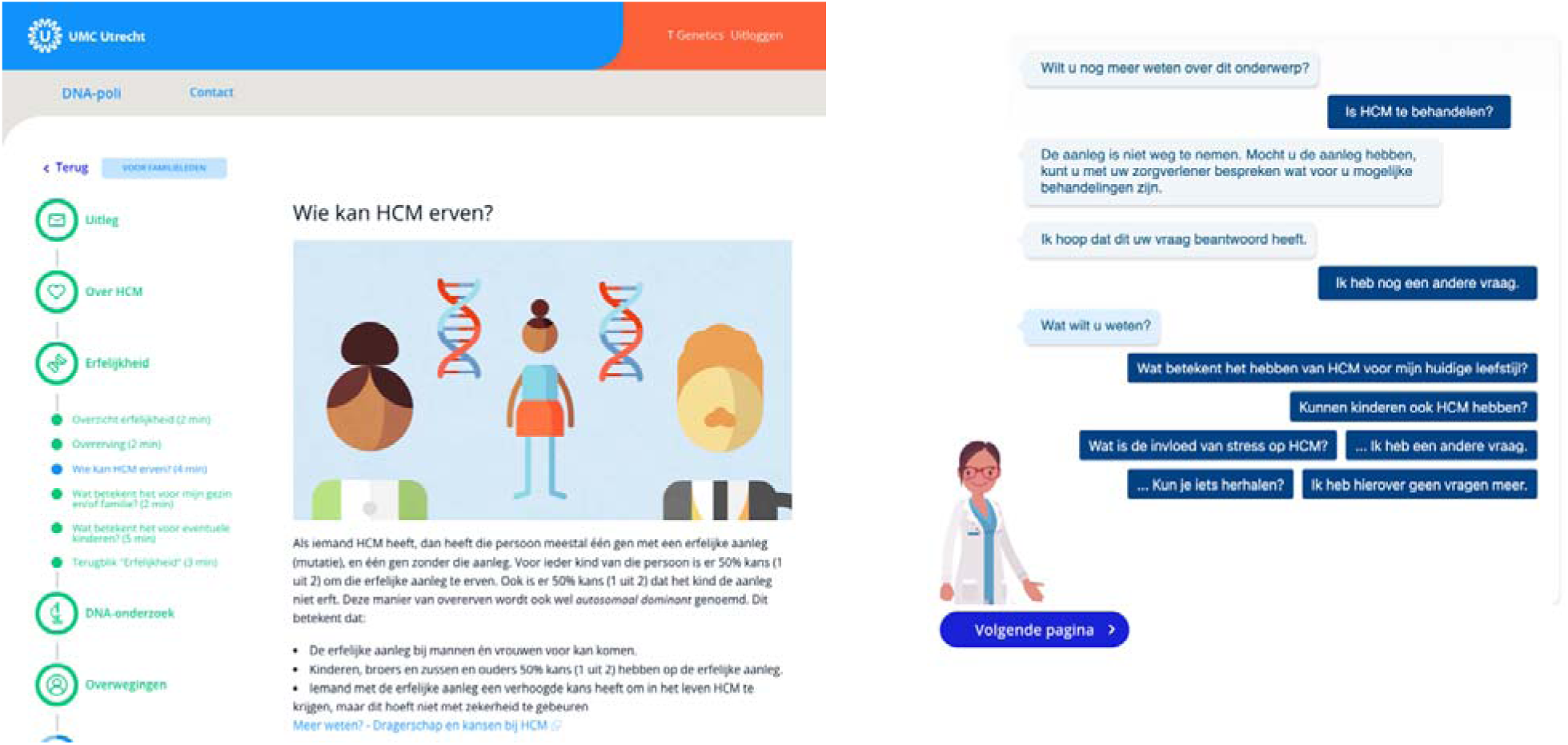
Visual impression of the design of DNA-poli. Figure 1a (left) portrays the user interface for at-risk relatives, showing different aspects of the genetic counselling pathway. Figure 1b (right) shows an impression of the virtual assistant “Anna” with a prescripted dialogue with an at-risk relative about HCM. HCM: hypertrophic cardiomyopathy.

If ARRs prefer genetic testing, or if they are in doubt, a mandatory remote telephone or video consultation with a genetic counsellor is scheduled within one week. If genetic testing is not preferred at that time, a referral for cardiac screening is provided, along with the option to consult a genetic counsellor. Those ARRs who opt for genetic testing will receive a blood sample kit via mail after the consultation. The ARR can present the kit at a local clinical laboratory for blood sample collection, after which the sample will be sent to the clinical genetics laboratory. After analysis of the familial variant, the return of results is provided online through DNA-poli or via telephone, if requested by the ARR. If applicable, follow-up for cardiac or further cascade genetic screening will be arranged through regular clinical care pathways. The full DNA-poli pathway is illustrated in Figure 2.

**Figure 2.**
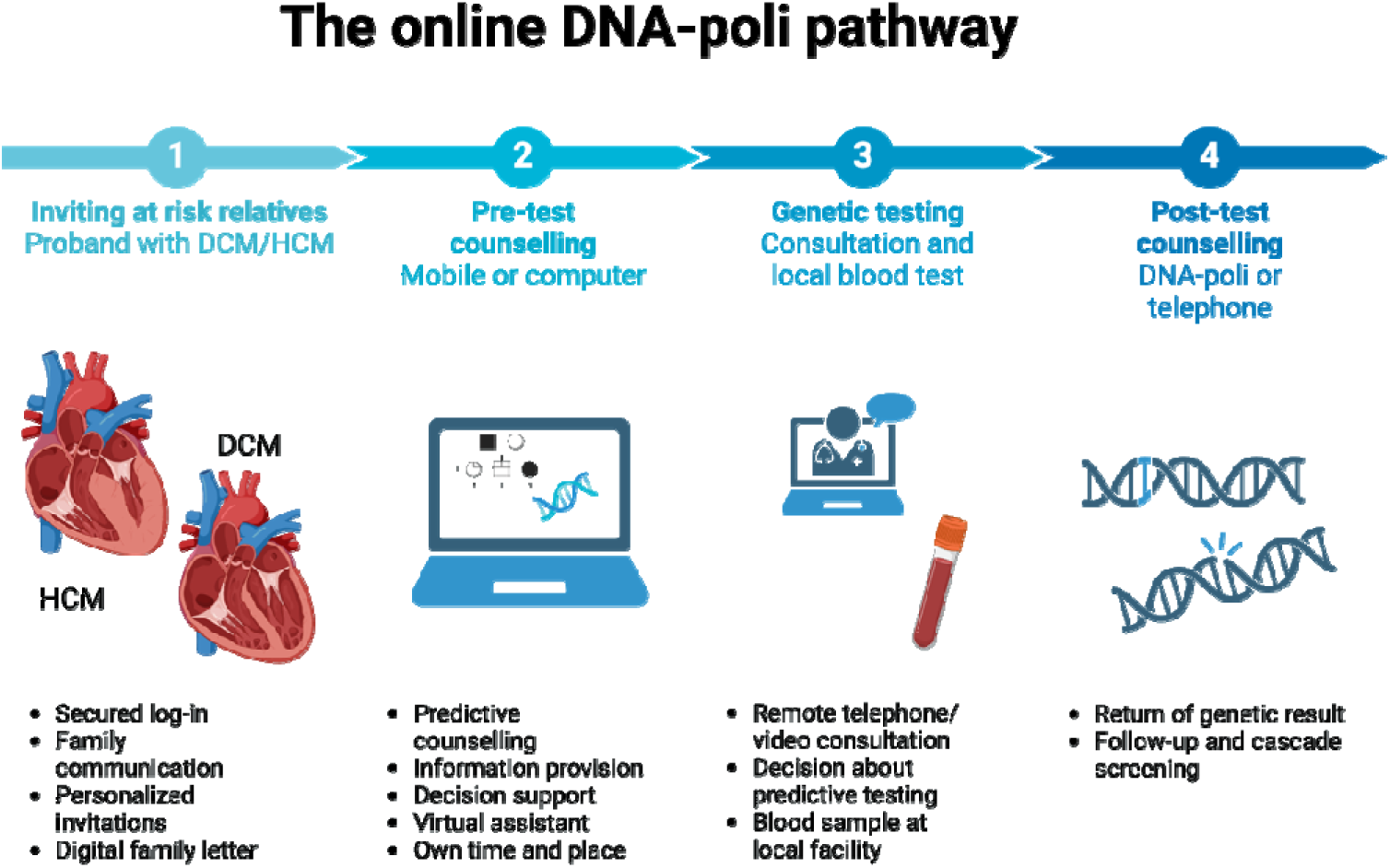
Overview of the DNA-poli journey for probands (Step 1) and their at-risk relatives (Step 2-4). DNA-poli supports family communication, pre- and post-test genetic counselling for families at risk of cardiogenetic disease. DCM: dilated cardiomyopathy, HCM: hypertrophic cardiomyopathy.

### Objectives

The primary aim of this RCT is to evaluate the uptake of cardiogenetic counselling six months after the proband’s genetic result disclosure with an extended follow-up of one year. In addition, experiences of probands, ARRs, and genetic HCPs will be collected and compared (DNA-poli versus standard clinical genetic care). Secondary objectives are focused on the quality of genetic counselling, including evaluating informed decision-making of ARRs, empowerment, and satisfaction in all stakeholder groups and in both arms of the trial.

## METHODS AND ANALYSIS

### Study design

This multicentre non-inferiority RCT with an unblinded parallel-group design will be conducted at six cardiogenetics departments of academic hospitals in the Netherlands (i.e., University Medical Centre Utrecht (UMCU), Amsterdam University Medical Centre (AUMC), University Medical Centre Groningen (UMCG), Radboud University Medical Centre (RadboudUMC) and Maastricht University Medical Centre (MUMC)). The effects of implementing the DNA-poli platform in pre- and post-test genetic counselling (intervention group) will be compared to current clinical genetic care (control group). Non-inferiority is reached if the primary outcome measures in the intervention group are comparable to or exceed the control group. Blinding of this study was not possible as participants in the intervention group are aware that they use a digital tool. This protocol has been drafted following the SPIRIT checklist [18].

### Participants

Three groups of stakeholders will be invited to participate: probands, ARRs, and HCPs. Probands are invited for study participation if they: (1) are the first family member to visit the outpatient clinic for the respective ICC, (2) are carriers of a likely pathogenic American College of Medical Genetics and Genomics (ACMG) Class 4 or pathogenic ACMG class 5 ICC related variant associated with HCM or DCM in one of the following genes with definitive gene-disease validity [19 ,20]: HCM: ACTC1, MYBPC3, MYH7, MYL2, MYL3, TNNC1, TNNI3, TNNT2, TPM1, or DCM: MYH7, TNNC1, TNNT2, TTN.

Also, probands (3) must be 18 years or older, (4) have sufficient digital skills, (5) have access to the internet and a mobile device or computer, (6) and must be proficient in Dutch to be eligible for inclusion. ARRs are eligible for inclusion if they meet criteria 3 to 6 listed above. Additional inclusion criteria for ARRs include that ARRs (1) are a first-degree relative of the proband or second-degree relative in case of a deceased first-degree relative, (2) do not present with clinically suspected HCM or DCM. Genetic HCPs fulfil inclusion criteria if they: (1) are employed by a genetics department in the Netherlands, and (2) are directly involved in counselling of ARRs for predictive cardiogenetic testing in the DNA-poli.

### Procedures

#### Recruitment and consent

Eligible probands will be consecutively invited to participate until the sample size is reached. Once consented to participate, probands will be followed for 12 months to assess the uptake of genetic counselling within each family. This time frame is chosen as previous studies have shown that the uptake of genetic counselling in ARRs increases most in the first six months and stabilises one-year post-disclosure of genetic test results in probands [9]. Probands are screened for eligibility by genetic HCPs and the executing researcher (ML) biweekly. Eligible probands will be informed about the RCT by their HCP during post-test counselling. If interested, the proband receives further study information and an informed consent form via email (using Castor Electronic Data Capture, New York, USA). Subsequently, the study team will contact the proband via telephone the next business day, to clarify the trial and to answer any remaining questions. Seven and 14 days after post-test counselling the proband is re-contacted by the study team to answer remaining questions and to explain study procedures. The proband is requested to decide on study participation within two weeks after receiving the invitation, otherwise, the proband is excluded, and cascade testing will continue as usual with a family letter. Subsequently, all included probands are asked to notify their ARRs about DNA-poli and the RCT and ask their permission to share personal contact details (name, e-mail address, telephone). Additional study information and informed consent forms are provided to ARRs either via Castor EDC by the study team after their intake consultation (control group) or automatically via DNA-poli after a first log-in (intervention group).

All HCPs involved in DNA-poli counselling will be trained by the executing researcher (ML) during a one-on-one training session and have access to the detailed standard-of-practice guidelines, which are updated when necessary. If a participant withdraws from the study, all data collected until that moment will be included in the analysis, unless the participant requests otherwise.

#### Randomisation

After providing consent, probands will be randomised to either the control or intervention group. Randomisation will be stratified by disease type (HCM or DCM) and participating centre. Castor EDC will be used for randomisation, with an allocation rate of 1:1. Subsequently, ARRs will be assigned to the same group as the proband they are related to. The proband and involved HCP will be informed via email about which group the proband is assigned to.

#### Control group

Probands in the control group will receive current clinical-genetic care at their local facility. They are asked to inform their ARRs, supported by a family letter which is posted by mail by the genetic HCP. This family letter includes a referral page so that ARRs can be referred for genetic counselling by their GP. After pre-test counselling, which is commonly done via telegenetics (i.e., video call or telephone consultation), the ARR decides whether or not to proceed with predictive DNA testing. DNA test results will be communicated by the HCP either via a telephone or video consult, or an in-person consultation. Afterward, each ARR receives a letter by mail summarising their care visit, including their decision regarding DNA testing, and if applicable, the test results and/or referral to a cardiology centre for clinical follow-up.

#### Intervention group

For individuals in the intervention group, DNA-poli will be used for family communication, pre-test, and if predictive DNA-testing is pursued, post-test genetic counselling (see also “Setting and intervention” section). First, probands receive an email with a link to access DNA-poli after securing two-factor authentication. Next, probands are asked to inform ARRs about the ICC diagnosis in the family and DNA-poli, as well as request their permission to add personal details to the platform. DNA- poli gives an overview of ARRs as discussed with the genetic HCP. Probands can fill out the personal characteristics (i.e., name, date of birth, email address, telephone number, and sex) before inviting them individually to DNA-poli. After two-factor authentication by email address and text message, ARRs can access a digital family letter in the DNA-poli platform and save a copy for printing. After reading study information and signing informed e-consent on DNA-poli, ARRs gain access to DNA-poli. They are automatically presented with either the DCM or HCM version of the pre-test counselling platform containing the subjects as described in the “Setting and intervention” section, depending on the ICC diagnosis in the proband. At the end of the online pre-test counselling, ARRs can indicate their initial preference regarding genetic testing, discuss their medical history with a virtual assistant, and, if desired, directly request a consultation with an HCP. This consultation (via telephone unless requested otherwise) is scheduled within one week. During the consultation, the ARR decides whether or not to proceed with DNA testing and how potential test results will be communicated (via the DNA-poli platform or telephone).

#### Multicentre coordination

The study will be coordinated by the UMC Utrecht research team. If the UMC Utrecht is not the primary care facility of the proband, this proband will be referred to the UMC Utrecht for follow-up of ARRs via DNA-poli, if randomised to the intervention group. The original centre remains in charge of clinical care for the proband involved.

### Patient and public involvement statement

Probands, relatives, patient representatives, and members of the general public were involved in the design and development of the DNA-poli prototype. Patients were not involved in identifying the research question or study design. Study results will be disseminated to study participants upon request. The authors will disseminate the study results via international and national conference presentations and presentations for patient societies.

### Measurement time points

Measures are collected at different time points using surveys, electronic health records, and DNA-poli output data, as displayed in Figure 3. All surveys will be sent via Castor EDC by the coordinating study team. First, probands will be asked to respond to one survey at 8 weeks (T1) post-disclosure of test results in both study groups. The survey will be administered regardless of whether all ARRs are invited. Second, ARRs are asked to participate in two surveys: one questionnaire after pre-test counselling (T1) and one eight weeks later, after receiving post-test counselling (T2). If post-test counselling is delayed, participants are requested to fill in the survey after post-test counselling.

**Figure 3.**
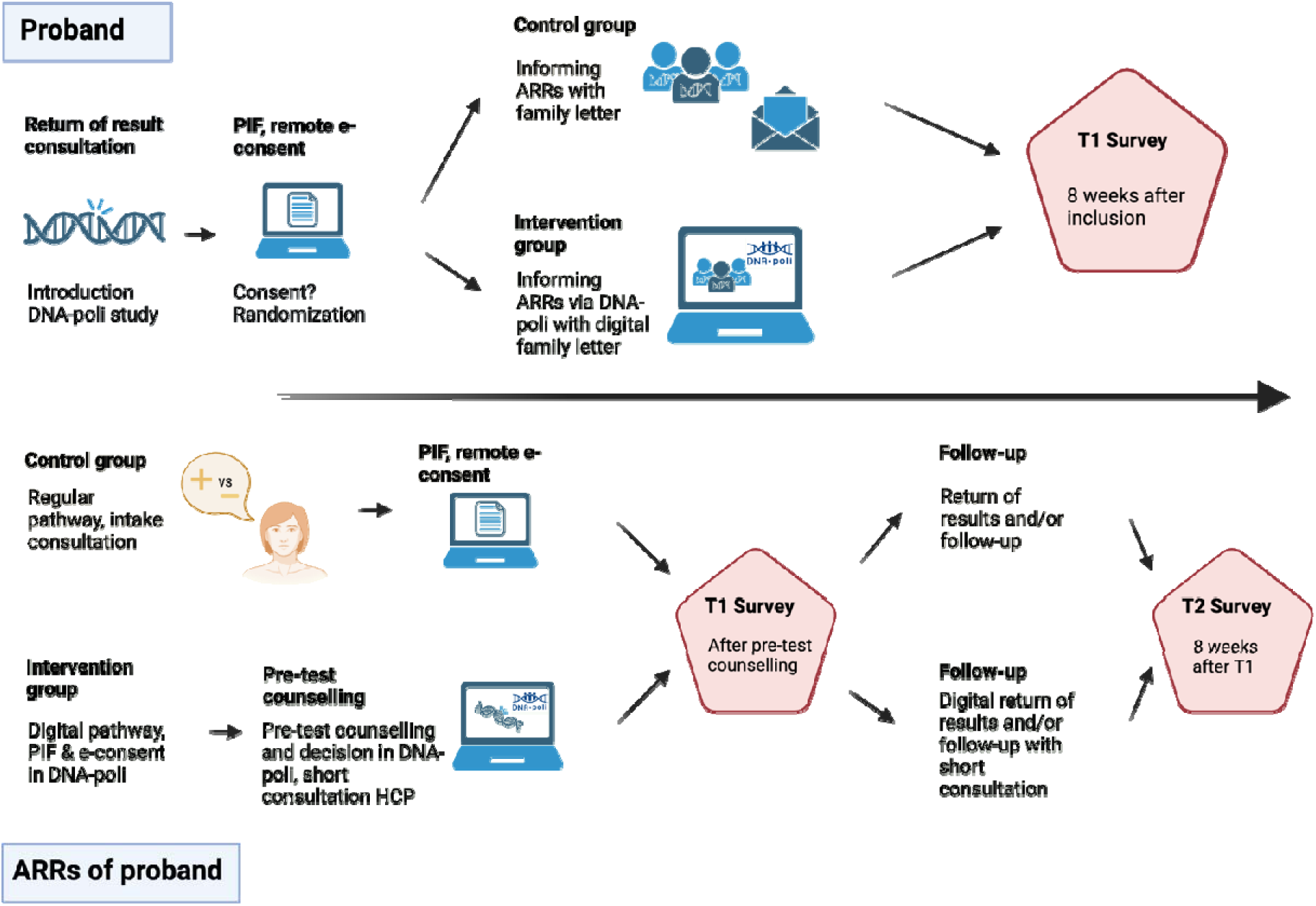
Overview of the DNA-poli trial pathway. On top the study process for probands is displayed, who receive one survey (T1) 8 weeks post-disclosure of test results. On the bottom, the study process for at-risk relatives (ARRs) of the proband is represented. In yellow, the control group is shown, who receive regular predictive counselling and genetic testing for the known variant in the proband. In green, the intervention group is shown, who receive online counselling in addition to a short consultation with an HCP. ARRs receive two surveys: T1 is after pre-test counselling and T2 is 8 weeks later. PIF: patient information folder, ARR: at-risk relative, HCP: healthcare professi nal.

Third, genetic HCPs are requested to fill out a survey once every three months during the course of this study. Participants will receive an automated reminder for completion two weeks after each survey via email if the survey is not completed in that time window. If surveys are not opened six weeks after the first invitation, one direct email will be sent to the participant by the study team to assess technical problems (e.g. survey invitation in a spam folder).

Electronic health record measures are collected throughout the study period, specifically, six months and one-year after the disclosure of proband’s test results. DNA-poli output data is collected during the use of the platform by the intervention group.

### Measures

Primary and secondary outcomemeasures collected in this study are summarised in Table 1

**Table 1:** Outcome measures Summarised outcome measures assessed in this study. For HCPs: T1 is after consultation with participants in the intervention group. For probands, T1 is 8 weeks post-disclosure of test results. For ARRs, T1 is after pre-test counselling and T2 is 8 weeks later. ARR: at-risk relative, HCP: healthcare professional, M: months, Y: year.

| Outcome measures | Data collection method | Data collection via | Participant and timepoint |
| --- | --- | --- | --- |
| <u>Primary</u> |  |  |  |
| Uptake, subjective | HCP assessment | Electronic health record | Proband, 6M&1Y post-disclosure of test results |
| Uptake, objective | Pedigree analysis | Electronic health record | Proband, 6M&1Y year post-disclosure of test results |
| Satisfaction: *Specific aspects of counselling* | Survey | Self-constructed survey | Proband (T1), ARR (T1, T2), HCP (T1) |
| Satisfaction: *General satisfaction* | Survey | Patient Satisfaction Questionnaire (PSQ), Dutch version [21] | ARR (T1, T2) |
| <u>Secondary</u> |  |  |  |
| Informed decision-making: *Decision* | (Preliminary) Decision about genetic testing | Electronic health record | ARR (T1) |
| Informed decision-making: *Decisional conflict* | Survey | Decisional Conflict Scale (DSC) [22], Dutch version [23] | ARR (T1, T2) |
| Informed decision-making: *Objective knowledge* | Survey | Knowledge questionnaire inherited oncological disease [24], adjusted version | ARR (T1, T2) |
| Empowerment | Survey | Genetic Counselling Outcome Scale-24 (GCOS-24) [25], Dutch version[26] | ARR (T1, T2) |
| Psychosocial impact | Survey | Shortened State-Trait Anxiety Inventory (STAI) [27] | Proband (T1), ARR (T1, T2) |
| Efficiency and patient contact: *Consultation and administration time* | HCP estimation of time investments | Electronic health record | HCP |
| Efficiency and patient contact: *Family cascade testing process* | Analysis of genetic care journey | Electronic health record, DNA-poli log data (intervention group) | ARR |
| Efficiency and patient contact: *Unscheduled consultations or contact* | Repeated data | Electronic health record, study e-mail | ARR |

#### Primary outcome measures

##### Uptake

In this study, uptake of genetic counselling is defined as the percentage of ARRs within one family that received or requested at least one pre-test consultation (control group) or finished pre-test counselling (DNA-poli and, if applicable, short pre-test consultation) with a genetic HCP within six months and one-year post-disclosure of proband’s test results. Similarly, uptake of genetic testing is defined as the percentage of ARRs within one family that continued with predictive testing within six months and one-year post-disclosure of proband test results. In collecting uptake data we will distinguish objective uptake, i.e. ARRs eligible for genetic testing based on pedigree analysis, and subjective uptake, i.e. ARRs eligible for participation according to the genetic HCP, in line with previous uptake studies [9].

The differences in uptake between the intervention and control group will be measured at two time-points (six months post-disclosure of proband test results with an extended follow-up of one year) in three ways: (1) the number of eligible ARRs logging in at the “DNA poli” and accessing the digital family letter (intervention group) or the number of ARRs referred for genetic counselling by the general practitioner, (2) the number of eligible ARRs attending pre-test genetic counselling (control group) or the number of ARRs consenting for DNA-poli and proceeding with pre-test counselling in DNA-poli and, if applicable, short pre-test consultation (intervention group), and (3) the number of eligible ARRs proceeding with predictive DNA-testing in both the control and intervention group. Per time-point, the number of ARRs will be compared to the total number of ARRs eligible for genetic counselling.

##### Satisfaction

Experiences with DNA-poli will be evaluated within each participant group. The satisfaction of participants with specific aspects of DNA-poli or regular counselling will be measured by a self-constructed questionnaire that will be administered in both groups. Questions focus on satisfaction with the usability, trustworthiness, and information density of the platform (intervention group) or counselling consultation (control group). The questionnaire consists of eight questions using a five-point Likert Scale (1 = ‘Totally disagree’ to 5 = ‘Totally agree’; range 5-40) and will be administered to ARRs at T1 and T2, to probands at T1 and in HCPs in all surveys. A score will be calculated by the sum of all items, where a high score indicates higher satisfaction. For ARRs, an adapted version of this questionnaire about the virtual assistant will also be provided, where DNA-poli is replaced by “virtual assistant Anna”.

To gain additional insights into general satisfaction with provided care, the Dutch version of the Patient Satisfaction Questionnaire (PSQ) will be added to the surveys for ARRs at T1 and T2 [21]. This questionnaire consists of 5 questions using a 10-point scale (1 = ‘Not at all’ to 10 = ‘A lot’; range 5-50). For the survey in the intervention group, the word “physician” will be replaced by “DNA-poli”, to align with participants’ experiences. Psychometric characteristics of the Dutch PSQ are considered satisfactory [21].

#### Secondary outcome measures

Secondary outcome measures will be collected using, electronic healthcare records and DNA-poli output data. Castor EDC will be used to administer the surveys and monitor survey progression. An overview of survey items is shown in Supplementary S1.

##### Informed decision-making

First, decision outcomes concerning the preliminary preference and final decision about genetic testing will be collected from electronic healthcare records. In addition, the Decisional Conflict Scale (DSC) questionnaire will be administered to ARRs at T1 and T2 [22 ,23]. The DSC measures whether participants feel uncertain, uninformed, unclear, and/or unsupported which could lead to ineffective decision-making. The DSC consists of 16 questions using a five-point Likert scale (1 = ‘Totally disagree’ to 5 = ‘Totally agree’). Scores range from 16 to 80, with a higher score indicating higher certainty about the decision made about predictive testing. Psychometric characteristics of the Dutch version of the DSC are considered sufficient [23]. In addition, objective knowledge is measured using an adapted version of a Dutch knowledge questionnaire that was developed for patients with inherited oncological disease in ARRs at T1 and T2 [24]. This questionnaire is considered suitable as it assesses knowledge about monogenic inheritance patterns, among others. In three questions about inheritance the terms “breast or ovarian cancer” are replaced by “heart disease”. Two remaining questions focusing on oncological aetiology are replaced by self-constructed similar questions about ICCs. In total, five knowledge questions are administered, which could be answered with ‘Yes’, ‘No’, or ‘I do not know’. A total correct knowledge score ranging from 0 to 5 is calculated, where a high score indicates a high knowledge level.

##### Empowerment

To assess empowerment after genetic counselling the Genetic Counselling Outcome Scale-24 (GCOS- 24) will be administered in ARRs at T1 and T2 [25]. The GCOS-24 covers themes related to experienced counselee empowerment, such as perceived personal and health control, mental state, satisfaction, and authenticity. In this study, the validated Dutch version of the GCOS-24 will be used [26]. In question 14, the phrase “clinical genetics service” was replaced with “DNA-poli” in the intervention group surveys, to align with participants’ experiences. The GCOS consists of 24 questions using a seven-point scale (1 = ‘Totally disagree’ to 7 = ‘Totally agree’; range 24-168). A high score indicates higher levels of empowerment. The psychometric characteristics of the Dutch GCOS are considered satisfactory [26].

##### Psychological impact

The shortened State-Trait Anxiety Inventory (STAI) questionnaire will be used to evaluate the psychological impact of genetic counselling among probands (T1) and ARRs (T1 and T2) [27]. The STAI consists of six questions on a four-point scale (1 = ‘Not at all’ to 4 = ‘A lot’; range 6-24). A high score indicates a more anxious participant. The psychometric characteristics of the shortened STAI are considered good [27].

##### Efficiency and patient contact

In each group, the efficiency of the counselling process from a HCPs perspective will be assessed by evaluating the time spent by the genetic HCP to prepare, counsel (consultation length), and complete follow-up administration for both intake and return of result consultations. Time spent (in minutes) will be registered in electronic patient files after each consultation by the HCP. For DNA-poli consultations without direct contact, only administration time will be registered. Efficiency of the counselling process for counselees is measured by comparing time spent on DNA-poli plus consultation time (intervention group) and consultation time alone (control group) in minutes. Time spent on DNA-poli will be estimated via log data from DNA-poli software. Efficiency of the cascade testing process will be assessed by evaluating the time between the proband test result, first intake consultation, and (if applicable) final consultation for at-risk relatives in both groups. Repeated data of unscheduled consultations with HCPs or contact moments with the study team will be documented in Castor EDC and electronic patient files.

### Participant characteristics

During T1, information about the sociodemographic and educational background of all probands and ARRs will be collected, including age, gender, highest completed education, living situation, parenthood and proband diagnosis at T1. Additionally, information is gathered about the age, gender, and type of HCP.

### (Digital) literacy

Digital literacy will be assessed at T1 for ARRs and probands by a shortened version of the Dutch eHealth Literacy scale in both the control and intervention groups [28]. This questionnaire consists of a list of five questions indicating an individual’s capability to evaluate electronically obtained health information and hereby assess digital literacy. The items are scored on a five-point Likert Scale (1 = ‘Totally disagree; to 5 = ‘Totally agree’; range 5-25). The higher the score, the more capable the individual is to evaluate health information. Psychometric characteristics of the eHealth Literacy Scale are considered sufficient [28 ,29]. To evaluate further health literacy, the ‘Functional Health Literacy’ and ‘Communicative Health Literacy’ items on the 3HL questionnaire will be administered [30]. The subscales consist of five items on a four-point Likert scale (1 = ‘Never’ to 4= ‘Often’; range 5-20).

These items indicate the capability of participants to obtain and understand medical information retrieved from a health organisation or by themselves. A lower score indicates a higher health literacy level. The psychometric characteristics of both subscales were assessed as good [30].

### Sample size calculation

Sample size calculation was based on the primary outcome of the uptake of genetic counselling. The study aims to illustrate an increased uptake of 15% points in the intervention group (mean uptake 56%, 3.05 ARRs per family counselled, estimated SD of 1.5) compared to the control group (mean uptake 41%, 2.00 ARRs per family counselled, estimated SD of 1.05). The control group mean is based on a previous study performed by Van den Heuvel et al. which showed the median pre-symptomatic ARRs eligible for pre-genetic counselling is 5.00 per family, with a range of 2 to 17 ARRs. Of these ARRs, 41.0% (N=295/717) attended genetic counselling in the first year, which indicates a mean of 2 relatives per family. A standard deviation of 1.5 was estimated. Assuming a power of 80%, a two-sided significance level of 5%, and an effect size d of 0.7 (mean intervention – mean control 3.05 – 2.00 = 1.05, difference / SD 1.05 / 1.50 = 0.7) the G*power calculation resulted in a total sample size of 253 individuals (68 probands, 170 ARRs, and 15 HCPs). It is expected that these individuals will be included in the trial within 21 months.

### Data analysis

All analyses will be performed in R statistics using RStudio. Descriptive statistics will be used to assess demographic information and efficiency measures. Continuous data will be tested for normality using the Kolmogorov-Smirnov test and graphical assessment. To assess differences in participant characteristics between randomisation groups, Mann-Whitney U tests for skewed continuous data and unpaired T-tests for normally distributed continuous data will be performed. Categorical variables will be presented as frequencies and their associated percentages, and the Chi-square test will be used to evaluate differences between groups.

To compare the intervention and control group on the primary outcome uptake of counselling, family-clustered logistic regression analysis will be performed, with relatives nested within families. In this logistic regression analysis, covariates will be included to control for confounding effects. For the primary outcome of patient satisfaction and secondary outcomes, independent sample T-tests or Mann-Whitney U tests will be used for continuous survey data. Chi-square tests will be used for categorical data. To assess data at different time measurement points, the paired T-test will be used for normal data and the Wilcoxon signed rank test for skewed data. The two measurement time points T1 and T2 in ARRs will be treated as nested within ARRs. A Bonferroni-corrected p-value of p<0.05 will be considered statistically significant. Multilevel mixed model analyses will be performed to assess the effect of randomisation, informed decision-making, and anxiety in ARRs and probands, adjusted for covariates.

## ETHICS AND DISSEMINATION

### Ethics

The study protocol was exempted from approval by the Research Ethics Committee NedMec because the Act of Medical Research Involving Human Subject (WMO) was not applicable (no. 23-066/C). The study protocol was also approved by local IRBs of the participating sites. We will obtain online informed consent from all participants. In case of any questions, the research team at the UMC Utrecht will answer questions via phone or e-mail. Participants will receive a copy of the informed consent form via DNA-poli or Castor EDC. All data collected for this study will be kept confidential.

Source data is stored at each participating cite while study data is stored in a secured research folder at the UMC Utrecht. There will be data sharing agreements between UMC Utrecht (UMCU) and each of the participating sites. The study will not have a data monitoring committee as we do not anticipate severe adverse effects, and it was not required for our study. To assure compliance with study protocols and ethical standards, the UMCU regularly conducts audits of research studies. Protocol amendments will be communicated to investigators and local IRBs of the participating cites and trial participants, If applicable.

### Dissemination

Alongside peer-reviewed journal publications, study findings will be shared with all stakeholders and during international and national conferences. Author eligibility will be based on ICMJE guidelines.

## TRIAL INFORMATION

The trial is registered with number NCT06431425 (ClinicalTrials.gov). Participant enrolment started in November 2023. The final proband is expected to be included in March 2025, and the final ARR in March 2026. This article describes the DNA-poli trial protocol version of March 2023. This trial is sponsored and coordinated by the University Medical Centre Utrecht (https://www.umcutrecht.nl). Study sponsor and funders had no role in study design.

## Data Availability

All data produced in the present study are available upon reasonable request to the authors

## ACKNOWLEDGEMENTS

We acknowledge Laura Yeates PhD. for her contribution to the final version of this paper. Figure 2 and 3 in this article were created in BioRender. van Tintelen, P. (2024) Figure 2: https://BioRender.com/a29u859, Figure 3 https://BioRender.com/p40c114.

## AUTHOR CONTRIBUTIONS

Conceptualization: ML, LH, PT, MS; Funding acquisition LH, PT, DH, MS; Writing original draft: JM, ML, LH; Review and editing manuscript: ML, LH, JM, PT, RK, TB, DH, MS; Supervision: ML, LH, PT.

## FUNDING STATEMENT

This work was supported by ZonMW/IMDI grant number 104021006 and the Dutch Heart Foundation grant number 2019B012.

## COMPETING INTEREST STATEMENT

Authors declare no conflicts of interest.

## DATA STATEMENT

The final trial dataset will be accessed by the principal investigator, members of the UMCU study team, and datamanager of the UMC Utrecht. All data produced in the present study are available upon reasonable request to the authors.

## SUPPLEMENTARY INFORMATION

Model informed consent forms, surveys, data management plan, and standards of practice are available in Dutch upon reasonable request.

## Notes

### Competing Interest Statement

The authors have declared no competing interest.

### Clinical Trial

NCT06431425

### Clinical Protocols

https://spirit-statement.org/

### Author Declarations

This study protocol was exempted from approval by the Medical Ethical Committee NedMec because the Act of Medical Research Involving Human Subject (WMO) was not applicable (no. 23-066/C).

## REFERENCES

1. Wilde AAM, Behr ER. Genetic testing for inherited cardiac disease. Nature Reviews Cardiology 2013;10(10):571–83. doi: 10.1038/nrcardio.2013.108

2. Lopes LR, Losi M-A, Sheikh N, et al. Association between common cardiovascular risk factors and clinical phenotype in patients with hypertrophic cardiomyopathy from the European Society of Cardiology (ESC) EurObservational Research Programme (EORP) Cardiomyopathy/Myocarditis registry. European Heart Journal - Quality of Care and Clinical Outcomes 2022;9(1):42–53. doi: 10.1093/ehjqcco/qcac006

3. Watkins H, Ashrafian H, Redwood C. Inherited Cardiomyopathies. New England Journal of Medicine 2011;364(17):1643-56. doi: 10.1056/nejmra0902923

4. Arbelo E, Protonotarios A, Gimeno JR, et al. 2023 ESC Guidelines for the management of cardiomyopathies. European Heart Journal 2023;44(37):3503-626. doi: 10.1093/eurheartj/ehad194

5. Tfelt-Hansen J, Garcia R, Albert C, et al. Risk stratification of sudden cardiac death: a review. Europace 2023;25(8) doi: 10.1093/europace/euad203

6. Van Den Heuvel LM, Hoedemaekers YM, Baas AF, et al. A tailored approach to informing relatives at risk of inherited cardiac conditions: results of a randomised controlled trial. European Journal of Human Genetics 2022;30(2):203–10. doi: 10.1038/s41431-021-00993-9

7. Heuvel LM, Smets EMA, Tintelen JP, et al. How to inform relatives at risk of hereditary diseases? A mixed-methods systematic review on patient attitudes. Journal of Genetic Counseling 2019;28(5):1042–58. doi: 10.1002/jgc4.1143

8. Van Der Roest WP, Pennings JM, Bakker M, et al. Family letters are an effective way to inform relatives about inherited cardiac disease. American Journal of Medical Genetics Part A 2009;149A(3):357-63. doi: 10.1002/ajmg.a.32672

9. Van Den Heuvel LM, Van Teijlingen MO, Van Der Roest W, et al. Long-Term Follow-Up Study on the Uptake of Genetic Counseling and Predictive DNA Testing in Inherited Cardiac Conditions. Circulation: Genomic and Precision Medicine 2020;13(5):524–30. doi: 10.1161/circgen.119.002803

10. Christiaans I, Birnie E, Bonsel GJ, et al. Uptake of genetic counselling and predictive DNA testing in hypertrophic cardiomyopathy. European Journal of Human Genetics 2008;16(10):1201–07. doi: 10.1038/ejhg.2008.92

11. Dragojlovic N, Borle K, Kopac N, et al. The composition and capacity of the clinical genetics workforce in high-income countries: a scoping review. Genetics in Medicine 2020;22(9):1437–49. doi: 10.1038/s41436-020-0825-2

12. Schmidlen T, Jones CL, Campbell-Salome G, et al. Use of a chatbot to increase uptake of cascade genetic testing. Journal of Genetic Counseling 2022 doi: 10.1002/JGC4.1592

13. Bombard Y, Ginsburg GS, Sturm AC, et al. Digital health-enabled genomics: Opportunities and challenges. The American Journal of Human Genetics 2022;109:1190–98. doi: 10.1016/j.ajhg.2022.05.001

14. Lee W, Shickh S, Assamad D, et al. Patient-facing digital tools for delivering genetic services: a systematic review. Journal of Medical Genetics 2023;60(1):1–10. doi: 10.1136/jmg-2022-108653

15. Christian S, Tagoe J, Delday L, et al. IMPACT webinars: Improving Patient Access to genetic Counselling and Testing using webinars—the Alberta experience with hypertrophic cardiomyopathy. Journal of Community Genetics 2022;13(1):81–89. doi: 10.1007/s12687-021-00564-x

16. Adam S, Birch PH, Coe RR, et al. Assessing an Interactive Online Tool to Support Parents’ Genomic Testing Decisions. Journal of Genetic Counseling 2019;28(1):10–17. doi: 10.1007/s10897-018-0281-1

17. Torr B, Jones C, Choi S, et al. A digital pathway for genetic testing in UK NHS patients with cancer: BRCA-DIRECT randomised study internal pilot. Journal of Medical Genetics 2022:jmedgenet-2022-. doi: 10.1136/jmg-2022-108655

18. Chan A-W, Tetzlaff JM, Altman DG, et al. SPIRIT 2013 Statement: Defining Standard Protocol Items for Clinical Trials. Annals of Internal Medicine 2013;158(3):200. doi: 10.7326/0003-4819-158-3-201302050-00583

19. Jordan E, Peterson L, Ai T, et al. Evidence-Based Assessment of Genes in Dilated Cardiomyopathy. Circulation 2021;144(1):7–19. doi: 10.1161/circulationaha.120.053033

20. Ingles J, Goldstein J, Thaxton C, et al. Evaluating the Clinical Validity of Hypertrophic Cardiomyopathy Genes. Circulation: Genomic and Precision Medicine 2019;12(2) doi: 10.1161/circgen.119.002460

21. Zandbelt LC, Smets EMA, Oort FJ, et al. Satisfaction with the outpatient encounter. Journal of General Internal Medicine 2004;19(11):1088–95. doi: 10.1111/j.1525-1497.2004.30420.x

22. Garvelink MM, Boland L, Klein K, et al. Decisional Conflict Scale Use over 20 Years: The Anniversary Review. Medical Decision Making 2019;39(4):301–14. doi: 10.1177/0272989x19851345

23. Koedoot NM, S.; Oosterveld, P.; Bakker, P.; de Graeff, A.; Nooy, M.; Varekamp, I.; de Haes, H. The decisional conflict scale: further validation in two samples of Dutch oncology patients. Patient education and counseling 2001;45(3):187–93. doi: doi: 10.1016/s0738-3991(01)00120-3

24. Pieterse AH, Ausems MGEM, Van Dulmen AM, et al. Initial cancer genetic counseling consultation: Change in counselees’ cognitions and anxiety, and association with addressing their needs and preferences. American Journal of Medical Genetics Part A 2005;137A(1):27-35. doi: 10.1002/ajmg.a.30839

25. Mcallister M, Wood A, Dunn G, et al. The Genetic Counseling Outcome Scale: a new patient-reported outcome measure for clinical genetics services. Clinical Genetics 2011;79(5):413–24. doi: 10.1111/j.1399-0004.2011.01636.x

26. Voorwinden JS, Plantinga M, Krijnen W, et al. A validated PROM in genetic counselling: the psychometric properties of the Dutch version of the Genetic Counselling Outcome Scale. European Journal of Human Genetics 2019;27(5):681–90. doi: 10.1038/s41431-018-0318-9

27. Van Der Bij AK, De Weerd S, Cikot RJLM, et al. Validation of the Dutch Short Form of the State Scale of the Spielberger State-Trait Anxiety Inventory: Considerations for Usage in Screening Outcomes. Public Health Genomics 2003;6(2):84–87. doi: 10.1159/000073003

28. Van Der Vaart R, Drossaert C. Development of the Digital Health Literacy Instrument: Measuring a Broad Spectrum of Health 1.0 and Health 2.0 Skills. Journal of Medical Internet Research 2017;19(1):e27. doi: 10.2196/jmir.6709

29. Norman CD, Skinner HA. eHEALS: The eHealth Literacy Scale. Journal of Medical Internet Research 2006;8(4):e27. doi: 10.2196/jmir.8.4.e27

30. van der Vaart RD, C. H.; Taal, E.; ten Klooster, P. M.; Hilderink-Koertshuis, R. T.; Klaase, J. M.; van de Laar, M. A. Validation of the Dutch functional, communicative and critical health literacy scales. Patient education and counseling 2012;89(1):82–88. doi: 10.1016/j.pec.2012.07.014

